# Dyslexia extends beyond reading difficulties: genetic links with social functioning and well-being

**DOI:** 10.64898/2026.10.30.26366900

**Authors:** Danielle Admiraal, Rada R. Veeneman, Else Eising, Simon E. Fisher

## Abstract

Dyslexia is a common neurodevelopmental condition characterized by persistent difficulties in reading and spelling. Although social and emotional challenges have been reported in individuals with dyslexia, the genetic relationships between dyslexia, social functioning, and well-being remain poorly understood. We combined phenotypic and genetic data from the Lifelines cohort with publicly available genome-wide association study (GWAS) summary statistics. Associations between dyslexia and social traits were examined using a multi-layered approach encompassing complementary methods, including phenotypic correlation analyses, polygenic score (PGS) analyses, linkage disequilibrium score regression (LDSC), and bidirectional two-sample Mendelian randomization (MR). Correlation analyses revealed significant phenotypic and genetic correlations between dyslexia and several social traits, including lower levels of life satisfaction, higher levels of loneliness, and less collegial support on the work floor. PGS analyses demonstrated that genetic liability to dyslexia predicted lower life satisfaction and reduced support from colleagues in the workplace, whereas none of the social-based PGSs predicted dyslexia. Bidirectional MR analyses provided evidence that genetic liability to dyslexia contributes to increased loneliness and social isolation, while no evidence was found for causal effects of social-interaction or well-being traits on dyslexia. Together, our results consistently supported a directional relationship from dyslexia toward social and well-being outcomes. In conclusion, our findings demonstrate that dyslexia shares genetic influences with social interaction and well-being traits and suggest that genetic liability for dyslexia contributes to adverse (psycho)social outcomes. These results extend the impact of dyslexia beyond literacy-related difficulties and highlight the importance of providing social and structural support to individuals with dyslexia throughout the lifespan.

## Introduction

In modern human society, written communication is used on a daily basis to facilitate social interaction. It serves multiple functions, including the transmission of information, expression of ideas, management of social relationships, articulation of needs, and transmission of knowledge and culture from one generation to the next. In essence, writing refers to the ability to encode linguistic structures into written symbols with orthographic rules, while reading refers to the ability to decode these symbols to reconstruct meaning. The level of skill for these complex processes differs among individuals, with more notable difficulties observed in individuals diagnosed with dyslexia(1). Dyslexia is a neurodevelopmental condition characterized by persistent difficulties in accurate and/or fluent word reading and spelling, despite adequate intelligence and educational opportunities(2). As reading and writing are fundamental skills for academic achievement and everyday functioning, understanding the nature and consequences of dyslexia is important.

Although written communication does not directly affect an individual’s verbal ability in social interaction, evidence suggests an indirect relationship between literacy skills and social well-being. For example, parents reported lower psychosocial functioning of their dyslexic children compared to typical readers already at 4 and 6 years of age, prior to their dyslexia diagnosis at age 8(3). Problems with psychosocial functioning and social interactions continue after entering school(3,4), as poor readers in the 3^rd^ grade are more likely to experience loneliness in the 5^th^ grade(5). In adulthood, reading ability among typically developing readers correlates positively with social-emotional skills (r=0.23; 95%CI=0.19, 0.28)(6). Taken together, these findings suggest that individuals who experience difficulties with written communication may be at increased risk of challenges in social interaction across the lifespan.

The level of difficulties and prevalence of dyslexia diagnoses is higher within families, which is explained in part by a hereditary component as well as the influence of shared environment. Reading outcomes are influenced by genetic components(7). However, an upbringing in which a child is frequently encouraged to engage in reading activities also improves overall reading ability(8). Genetic contributions to reading outcomes were first demonstrated in twin studies, where the heritability of a trait is deduced by using the difference in genetic similarity between mono- and dizygotic twins, while taking the shared environment to be constant. The largest twin-based meta-analysis on reading ability to date estimated heritability for general reading (h^2^_twin_=66%), letter-word knowledge (h^2^_twin_=62%), phonological decoding (h^2^_twin_=68%), reading comprehension (h^2^_twin_=68%), phonological awareness (h^2^_twin_=52%), and spelling (h^2^_twin_=80%)(9).

Large-scale genotyping efforts enable researchers to quantify relatedness among unrelated individuals from their genetic information, giving opportunities to estimate heritability of complex traits using single-nucleotide polymorphism (SNP) data. The regression of SNP data with phenotypic scores forms the basis of genome-wide association studies (GWAS)(10) that are used to identify DNA variants involved in heritable traits, and this approach has been applied to various reading-related traits. Doust et al. (2022) performed a GWAS on self-reported dyslexia, which was conducted in data of the direct-to-consumer genotyping company 23andMe (n=1,138,870 with n_cases_=51,800), and identified 42 significantly associated loci, with an estimated SNP-based heritability (h^2^_SNP_) of 0.18 (SE=0.008)(11). Eising et al. (2022) targeted five reading- and language-related skills in up to 34,000 people within the GenLang consortium and identified heritability for word reading (h^2^_SNP_ =0.19; SE=0.021), nonword reading (h^2^_SNP_ =0.26; SE=0.028), spelling (h^2^_SNP_ =0.23; SE=0.031), phoneme awareness (h^2^_SNP_ =0.25; SE=0.040), and nonword repetition(h^2^_SNP_ =0.13; SE=0.039)(12). The efforts of these large-scale GWAS studies have provided valuable data for further investigation of genetic associations with related outcomes, such as social interaction and well-being.

Few studies have investigated the shared genetic architecture between reading ability and social interaction experiences. Indications of genetic overlap have been provided by Doust et al. (2022), as analyses of their GWAS summary statistics yielded a significant genetic correlation between dyslexia and loneliness (r_g_=0.24)(11), while Eising et al. (2022) found a positive correlation between the combined reading-traits that they studied and friendship satisfaction (r_g_=0.15; SE=0.058), as well as a negative relationship with loneliness (r_g_=-0.24; SE=0.048)(12). However, genetic correlations between reading ability and a broader set of social interactive traits have not yet been investigated, nor has the power of genetic influences on reading ability as predictors of social outcomes.

Here, we investigated the shared genetic architecture and predictive power of dyslexia with respect to a range of social interactive traits. We used published GWAS summary statistics of dyslexia(11) and several social-interaction traits(13–16) combined with quantitative trait and self-report data of similar traits and SNP data in the Lifelines(17) cohort; a multi-disciplinary prospective population-based cohort study of individuals living in the North of the Netherlands. With these data, we i) assessed the relations between social-interactive traits in the Lifelines cohort using psychological network analysis, ii) investigated the shared genetic architecture between dyslexia and publicly available GWAS summary statistics representative of social interaction and well-being, iii) used polygenic score analysis to investigate the predictive power of genetic disposition to dyslexia for social-interactive outcomes and vice versa, and iv) tested both directions of causality for SNPs robustly associated with dyslexia and social interactive outcomes using two-sample mendelian randomization analysis. Together, these findings provide a comprehensive overview of the relationship between genetic architectures underlying dyslexia and social well-being.

## Results

### Psychological network-based phenotype selection

Data for this study were derived from the Lifelines(17) cohort; a multi-disciplinary prospective population-based cohort study of individuals living in the North of the Netherlands. Data on three reading-based items, obtained in 2022, were available for 31,131 to 33,859 individuals (Table 1). Thirteen (sub) scales representative of social interactive experiences were collected in two waves between 2012 and 2019 with a total of 54,264 to 123,833 participants (Table 2). Descriptives for all traits are reported in Supplementary Table 1.

**Table 1:**
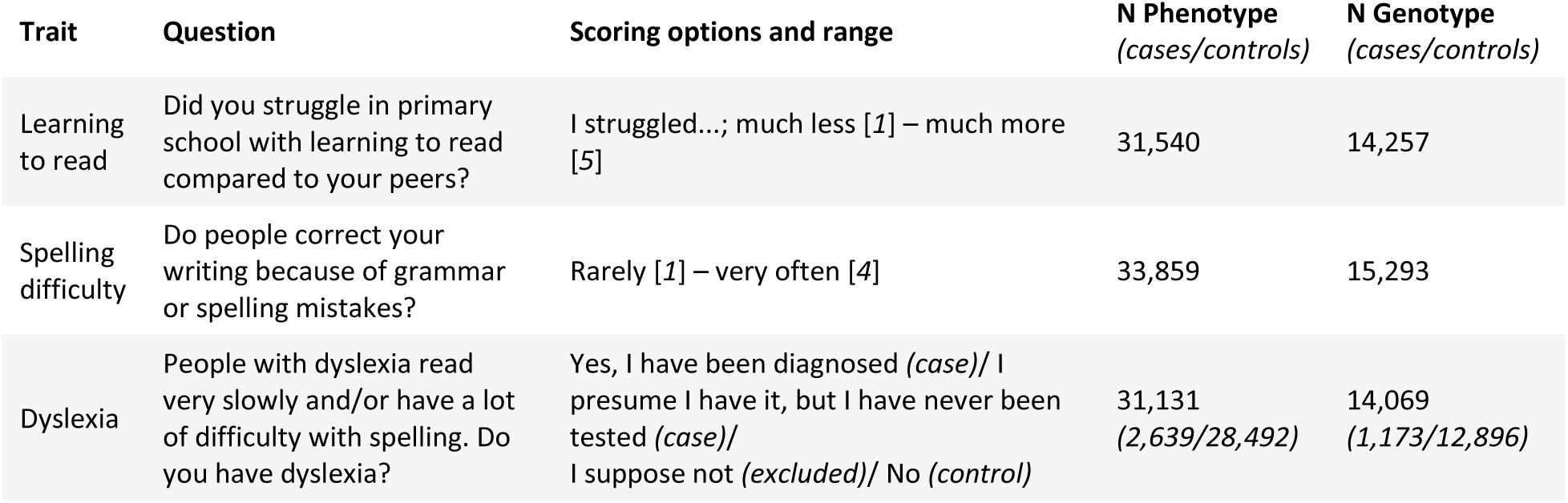
Overview of reading-based traits used in this study. The survey questions and scoring options are shown, along with the phenotype- and genotype-based sample sizes.

**Table 2:** Overview of social survey-based traits used for this study. The survey names and their respective scales are shown, along with the scoring ranges. Surveys that did not have any lower-level scales are indicated with n.a. (not available), and lower scores indicate less and/or lower social-interactive/well-being experiences. Phenotype- and genotype-based sample sizes are given.

| Survey | Scales | Scoring range | N Phenotype<br>*(cases/controls)* | N Genotype<br>*(cases/controls)* |
|---|---|---|---|---|
| Loneliness (LS) | *n.a.* | *Case/control* | 61,806<br>*(17,158/44,648)* | 26,707<br>*(7,061/19,646)* |
| Self-Management Ability Scale (SMAS) | Initiative taking | 0-100 | 91,433 | 40,549 |
|  | Investment behaviour |  |  |  |
|  | Resources availability |  |  |  |
|  | Multifunctional activities |  |  |  |
|  | Positive mind-state |  |  |  |
|  | Self-efficacy beliefs |  |  |  |
| Copenhagen Psychosocial Questionnaire (COPSOQ) | Social support colleagues | 0-100 | 54,264 | 24,308 |
|  | Social support supervisor |  | 59,673 | 26,753 |
| Satisfaction with Life Scale (SWLS) | *n.a.* | 5-35 | 61,848 | 26,705 |
| Social Production Function Instrument for the Level of well-being (SPFIL) | Affection | 0-100 | 123,833 | 55,743 |
|  | Confirmation |  |  |  |
|  | Status |  |  |  |

We calculated phenotypic correlations between reading ability and social interaction in Lifelines(17) (Suppl. Table 2). Correlations between the reading-based items were moderate and positive (*r*=0.40-0.45; *p*<2.2E-16), whereas the associations between reading- and social-based traits were low, yet significant, with correlations ranging from 0.016 between dyslexia and SMAS_multifunctionality (*p*<9.12E-3), to 0.128 between learning to read and SMAS_investment (*p*<2.2E-16). After Bonferroni correction (*p_Bonferroni_-*=0.05/120 tests = 4.17E-4), only 3 out of 91 tests between the reading- and social-based traits did not reach significance. Thus, the associations between reported reading skills and social-traits in Lifelines data were robust.

The number of scales for the social-based surveys differed. For example, the loneliness survey produced a single final score, and the SMAS survey yielded six scores for the six sub-scales. We reasoned that surveys with multiple scales were more prone to type I error, potentially leading to more biased results for surveys with multiple scales, compared to those that only consist of a single score. Therefore, for our genetic analyses, we selected one scale per social survey, and one item for the reading-based traits. First, we wanted to ensure that the selected traits could (partially) be explained by genome-wide SNP variance, estimating h^2^_SNP_ for all traits using GCTA-GREML(18) (Suppl. Figure 1). Estimates for the reading-related traits were low (h^2^_SNP_=0.13-0.16; *p*<1.02E-06) and social-trait estimates were even lower, yet still significant (h^2^_SNP_=0.04-0.09; *p*<5.00E-03). Second, we selected the sub-scales that most strongly represented their respective survey through psychological network analyses, which models the statistical relationships between two variables while conditioning on all other variable-relationships in the dataset, resulting in partial correlation coefficients(19,20). The network represents a complex structure of all traits (nodes) and their partial statistical relationships (edges). A mixed graphical model(21,22) approach was conducted in order to allow modelling for various data distributions.

The social-based network (Figure 1A) consisted of thirteen traits, distributed over 5 different surveys. All scales clustered together within their respective survey, meaning that after accounting for all correlations across the network, a large part of the shared variance across survey-specific sub-scales remained. Even though there are correlations between different survey scales, the smaller edges between sub-scales from different surveys in network show that the surveys are fairly distinct from one another, measuring different social-interactive aspects. For the surveys with multiple sub-scales, we selected the sub-scale with the highest strength index S_i_, which captures the sum of partial correlations it is connected with, for follow-up analyses. Within the social-based network, we thus selected SMAS self-efficacy (*S* = 1.23), SPFIL confirmation (*S* = 1.05), and COPSOQ colleagues (*S* = 0.77). In addition, we included LS (*S* = 1.27) and SWLS (*S* = 0.79) as these were made up of a single scale. The dyslexia item was the strongest driver in the reading-based network (*S* = 1.84) (Figure 1B), and was thus selected to represent reading ability in follow-up analyses.

**Figure 1:**
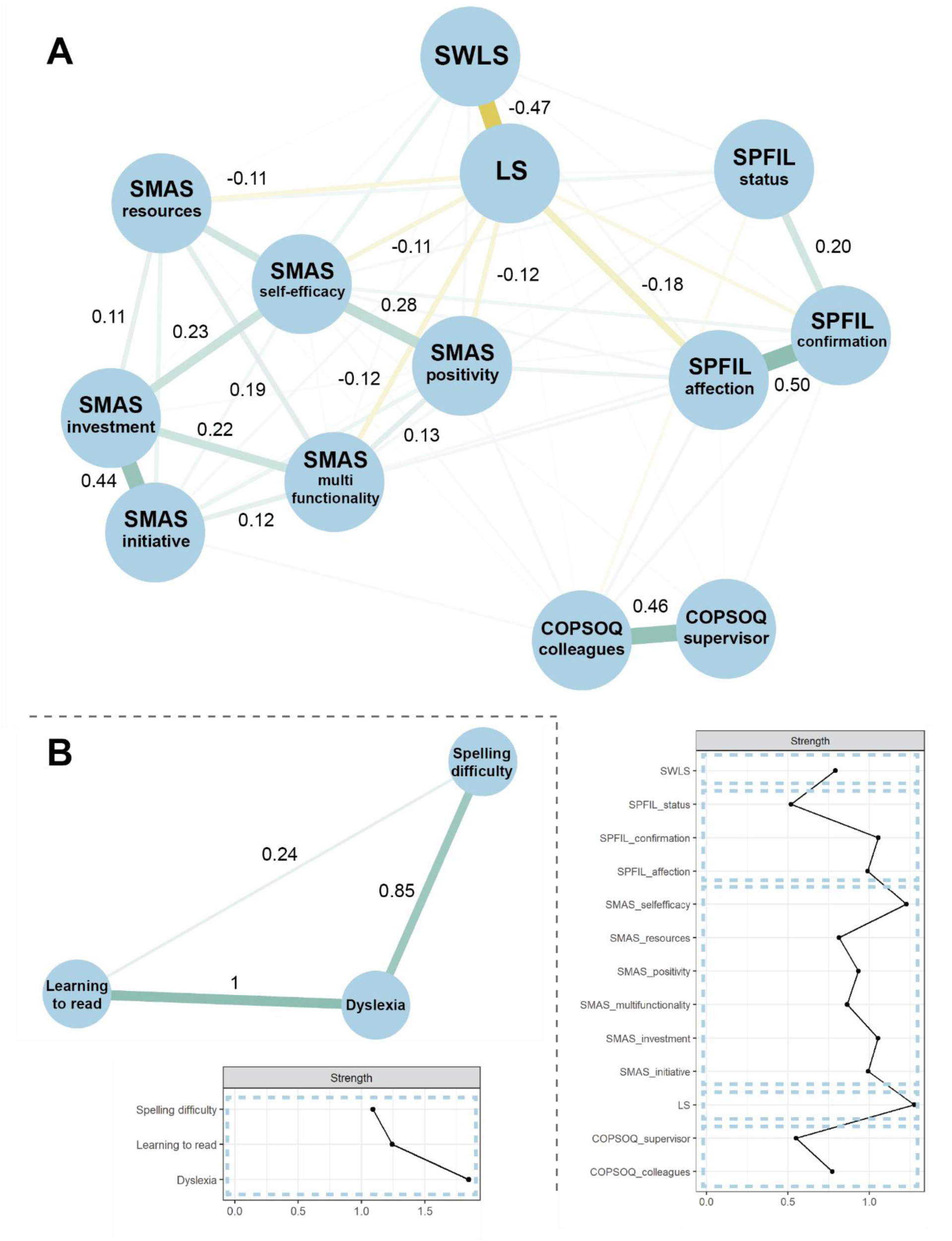
Psychological network analyses across (**A**) social-based (sub-)scales, and (**B**) reading traits. The network only displays edge values with strength index S ≥ 0.1 and p<0.05 (p-values based on a case-drop test with N_tests_=1,000). The network-associated strength indices of the respective nodes (Sᵢ) are presented in the line plot. Sub-scales and items belonging to the same instrument/survey are grouped by the dotted lines. The correlation stability coefficient (CS)-strength of both networks was estimated as CS_strength_=0.75.

### Genetic disposition for dyslexia predicts the amount of help received from colleagues on the work floor and overall life satisfaction

For maximal power, we took advantage of independent GWAS summary statistics available from the largest studies of these traits so far to calculate genetic correlations (r_g_) between social-interaction outcomes and dyslexia, using Linkage Disequilibrium Score Regression (LDSC)(23,24). We selected GWAS summary statistics for self-reported diagnoses of dyslexia (n=1,138,870 with n_cases_=51,800)(11), as well as social-based GWAS summary statistics for i) subjective well-being (n=192,994)(16), ii) levels of sociability (n=342,461)(14), iii) the experience of social trust towards strangers (n=33,882)(25), iv) the frequency of social interaction defined as social isolation (n=449,609)(15), and v) personal experience of loneliness combined with the frequency of social interaction defined as loneliness (n=452,302)(13) (Suppl. Table 5). Two social-based traits showed modest but significant genetic correlations with dyslexia after Bonferroni correction (*p*_Bonferroni_=0.05/5 tests=0.01), with r_g_=0.21 (*p*=5.71E-14) for social isolation, and r_g_=0.23 (*p*=1.43E-18) for loneliness.

Next, we used polygenic scores (PGS) to further investigate genetic overlaps between dyslexia and social-interaction outcomes. Because the heritability estimates for the social-based traits and their genetic correlations with dyslexia were low to modest (Suppl. Figure 1 and Suppl. Table 5), power calculations were run to investigate whether there was sufficient power for the PGS analyses. Following the power-prediction framework by Pasaniuc and Price et al. (2017)(26) (see Methods for more details), the dyslexia GWAS summary statistics combined with the Lifelines sample sizes had >80% power to identify association representing genetic correlations of r_g_>0.02 (Suppl. Figure 2). PGS based on the GWAS summary statistics for loneliness, sociability, and social isolation each had 80% power to predict variance in dyslexia representative of r_g_>0.03, and subjective well-being had 80% power to identify an association representing a genetic correlation of r_g_>0.06 (Suppl. Figure 3A). Only the PGS for social trust had insufficient power to explain enough variation in dyslexia and was therefore excluded from the PGS analyses.

We first ensured that the calculated PGSs could predict trait outcomes within their own category. PGS_dyslexia_ significantly predicted 5.6% of dyslexia variance in Lifelines (OR = 1.9; 95% CI = 1.8,2.2; *p*_OR_=4.1E-36) (Suppl. Figure 4A). PGSs based on all four social-based summary statistics significantly predicted all five social Lifelines traits, with explained variance ranging between 0.06-0.51% (2.9E-22<*p*_β_<0.014) (Suppl. Figure 4B). Next, we tested PGS_dyslexia_ for prediction of the social traits in Lifelines (Figure 2A, Suppl. Table 6). The dyslexia scores robustly explained variance for help from colleagues (IncR^2^ = 0.07%; β = −0.48 [95% CI = −0.13, −0.83], *p*_β_=7.36E-3) and life satisfaction (IncR^2^ = 0.11%; β = −0.22 [95% CI = −0.09, −0.36], *p*_β_=1.22E-3), with higher PGS_dyslexia_ being associated with lower levels of social-interactive outcomes. The remaining traits, social self-efficacy, social confirmation and loneliness, did not show significant relationships with PGS_dyslexia_. Lastly, we investigated whether genetic disposition for social-based traits could predict dyslexia (Figure 2B). However, none of the PGSs for social isolation(15), sociability(14), loneliness(13), and subjective well-being(16) explained any variance for dyslexia in the Lifelines sample.

**Figure 2:**
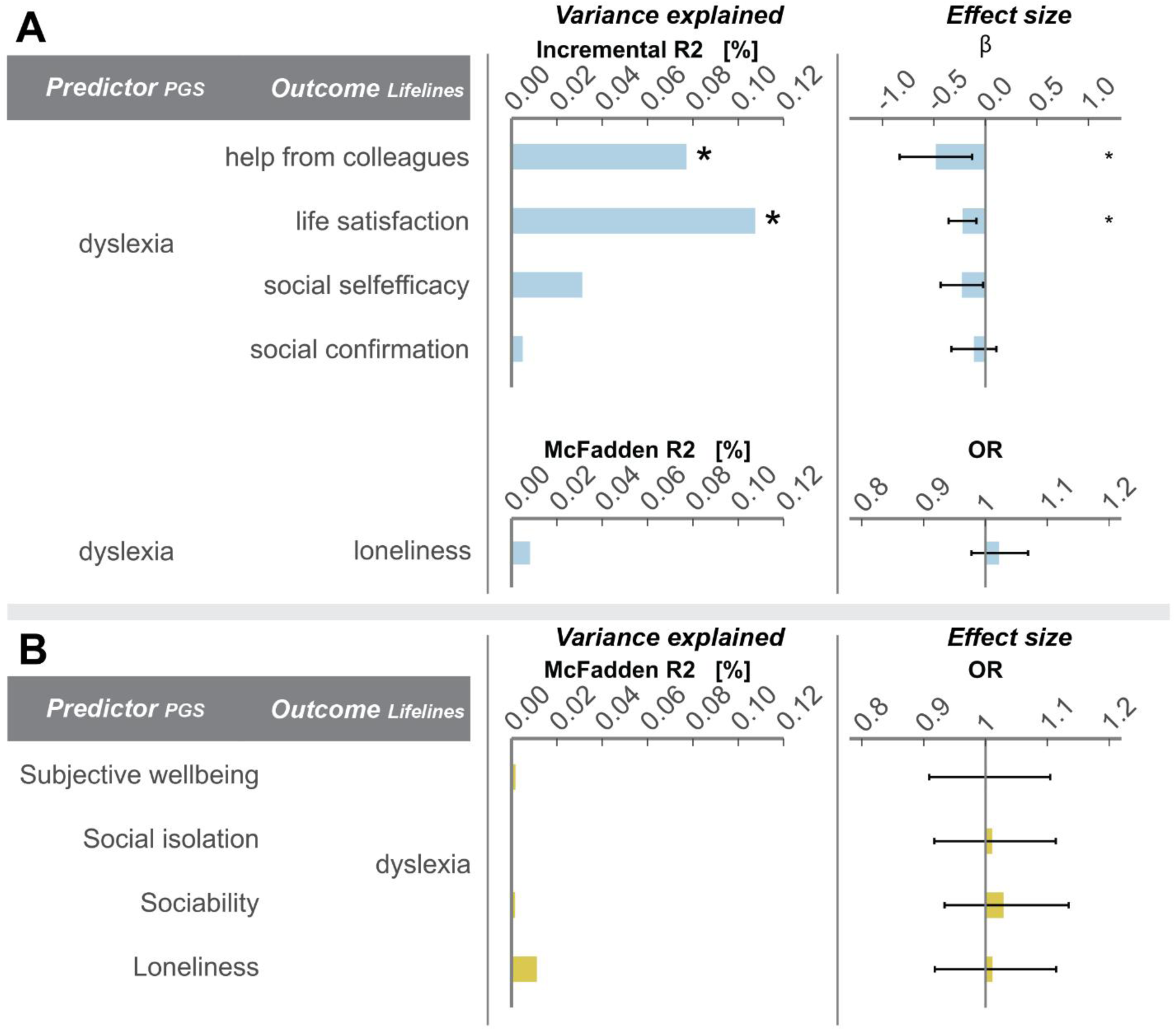
(**A**) PGS for dyslexia predicting social-based measures, and (**B**) PGS for various social interactive traits predicting dyslexia. The first column represents the predictor (GWAS-based) and outcome (Lifelines data) traits. The second column displays the explained variance for both continuous (Incremental R^2^) and binary (McFadden R^2^) outcomes. **\*** represents estimates that are significant after multiple testing correction (figure A = 5 tests for PGS_dyslexia_ → social outcome, gives p_bonferroni_=0.05/5=0.01; figure B = 4 tests for PGS_social_ → dyslexia, gives p_bonferroni_=0.05/4=0.0125). The third column shows the effect sizes, again for continuous (beta [β]) and binary (odds-ratio [OR]) outcomes, with error bars representative of 95% confidence intervals. Predictor GWAS summary statistics: Dyslexia by Doust et al. (2022)(11); Loneliness by Day et al. (2018)(13); Sociability by Bralten et al. (2021)(14); Social isolation by Socrates et al. (2024)(15); Subjective well-being by Okbay et al. (2016)(16).

We observed a skewed distribution of the incidence of dyslexia self-report across age in Lifelines (Suppl. Figure 5A). This might be explained by a change in the national protocol for diagnosing dyslexia in 2006 in the Netherlands, leading to policy changes and a rise in dyslexia diagnoses due to increased attention on this trait in schools(27). As none of the social PGSs could account for dyslexia variance in the sample, we were concerned that true genetic effects might have been masked by phenotypic noise as a result of less restrictive dyslexia diagnoses in the younger subsample due to societal pressures. We therefore further investigated predictive behaviour of the PGSs for dyslexia and of social isolation, the latter having the largest power to identify an association with dyslexia, for the two other reading-based traits available in Lifelines: spelling difficulty and learning to read (Suppl. Figure 5B). PGS_dyslexia_ explained less variance in spelling difficulty and learning to read than in the dyslexia outcome, even though results were still significant and robust (spelling difficulty, IncR^2^ = 2.0%, β = 0.08 [95% CI = 0.07, 0.10], *p*_β_=3.4E-27; learning to read, IncR^2^ = 1.8%; β = 0.14 [95% CI = 0.12, 0.16], *p*_β_=1.5E-23) (Suppl. Table 6). PGS_isolation_ could not predict any of the available reading-based outcomes (spelling difficulty IncR^2^ = 5.9E-3%, β = 4.54E-3 [95% CI = −1.05E-2, 1.96E-2], *p*_β_=5.54E-1; learning to read IncR^2^ = 4.9E-3%; β = 7.12E-3 [95% CI = −1.97E-2, 3.4E-2], *p*_β_=6.03E-1) (Suppl. Table 6). Therefore, changes in the national diagnostic protocol do not seem to have caused the lack of associations between the social PGSs and dyslexia.

### Causal implications for dyslexia towards social isolation and loneliness

To further investigate potential causal effects between dyslexia and social interaction, we performed mendelian randomization analyses using the dyslexia and social-based GWAS summary statistics(28,29). Analyses were performed with SNPs that reached genome-wide significance in the exposure GWAS (*p*<5E-8) as instrumental variables, after which we investigated their effect in the outcome GWAS summary statistics using five methods, of which the Inverse Variance Weighted (IVW) method was used as the main method, and the MR Egger, Simple mode, Weighted median, and Weighted mode were used as sensitivity analyses.

First, we used the dyslexia GWAS as an instrumental variable to investigate the causal relationship between dyslexia and the social-based GWASs (Suppl. Figure 6A, Suppl. Table 7). There was strong evidence that dyslexia predicted social isolation (β_IWV_ = 0.05; *p* = 8.3E-6) and loneliness (β_IWV_ = 0.04; *p* = 1.2E-5), even after Bonferroni correction (*p_Bonferroni_* = 0.05/4 tests = 0.0125). The Weighted median provided further support for a causal effect of dyslexia on loneliness (β_Weighted median_ = 0.03; *p* = 8.8E-3). While the Weighted Mode and Simple Mode did confirm the direction of effect size, they did not reach statistical significance to confirm the IVW (see Methods for more details on the sensitivity analyses). The Cochran Q test showed no evidence of heterogeneity among the instrumental SNPs for dyslexia on loneliness, subjective well-being, and social isolation (*p_Cochran Q_* > 0.08) (Suppl. Table 7). This was further supported by the leave-one-out analysis, which showed that excluding individual instrumental SNPs did not lead to a change in the IVW effect estimate (Suppl. Figure 7). Next, we investigated the causal effect of social-traits on dyslexia outcomes (Suppl. Figure 6A, Suppl. Table 7), but found no evidence for causal effects. Secondary analyses, with instrumental SNP variables that were selected based on a suggestive GWAS *p*-value threshold (*p*<1E-6), yielded highly similar results to the primary analyses (Suppl. Figure 6B, Suppl. Table 7). Social isolation and loneliness were predicted by the dyslexia SNPs, yet none of the social-instrumental variables predicted dyslexia, even with higher numbers of instrumental SNPs, suggesting that our results are robust. These findings show that i) there are indications that dyslexia-associated SNPs may be causal for loneliness and social isolation, and ii) the SNPs associated with sociability, social isolation, loneliness, subjective well-being, and social trust are not causal for dyslexia.

## Discussion

We investigated the phenotypic and genetic relations between dyslexia and social interaction/well-being traits using published GWAS summary statistics and direct phenotype and genotype data from the Lifelines cohort. When considered collectively, the phenotypic correlations, PGS, genetic correlations, and MR results provided a nuanced picture of the relationship between dyslexia and social functioning. First, we identified low yet significant phenotypic correlations between three reading traits and 13 measures of social interaction difficulties in adults. With 87 out of 91 phenotypic correlations surviving Bonferroni correction (*p_Bonferroni_*=0.05/91 tests=5.50E-4), we considered our phenotypic results robust. Previous studies have reported social interaction difficulties in children with dyslexia prior to entering school(3), as well as during the schooling period(5,6), yet our results show that also adults with dyslexia experience more challenges in their daily social interaction. Next, we examined genetic correlations between dyslexia and social traits, observing significant positive correlations with loneliness and social isolation. Previous studies reported similar findings, with genetic correlations between dyslexia and loneliness (r_g_=0.24; SE=0.034)(11) and between reading ability and friendship satisfaction (r_g_=0.15; SE=0.058) and loneliness (r_g_=-0.24; SE=0.048)(12). Higher PGS for dyslexia predicted overall lower life satisfaction and less support received from colleagues, whereas none of the social-trait PGSs predicted dyslexia. Notably, the strongest PGS association of dyslexia was observed for workplace support from colleagues. These work environment-specific experiences for individuals with dyslexia have been reported before, as they experience more challenges in work confidence(30) and social relationships with co-workers(31). Lastly, MR analyses showed the same directional pattern, with evidence for dyslexia-related variants affecting loneliness and social isolation but no evidence that the social-based SNP-variants have causal effects on dyslexia.

Across PGS and MR analyses, our results suggested a directional relationship in which genetic disposition to dyslexia influences social outcomes rather than the reverse. Evidence for this directionality was strongest for loneliness and social isolation. Other studies also found correlations between dyslexia and heightened levels of self-perceived anxiety, depressive symptoms, and lower self-esteem(32–34). The International Dyslexia Association(35) describe how reading difficulties in day-to-day life, in spite of continuous efforts to prevent writing and reading mistakes, can give a heightened sense of anxiety and frustration. Furthermore, comparisons with typical readers in school, who can easily resolve reading and writing mistakes with practice, can lead to less confidence and lower levels of emotional well-being in individuals with dyslexia, with effects throughout the lifespan during schooling, studies, and working life. This description fits with our findings, and would explain why the underlying genetics for dyslexia may be causal towards social-interactive and well-being outcomes, and not the other way around.

To our knowledge, this is the most comprehensive investigation to date into the shared genetics underlying dyslexia and social well-being. A study with a comparable PGS framework and focus examined developmental language disorder (DLD) in relation to common psychiatric disorders, including major depressive disorder, anxiety disorder, and ADHD(36). That study found that individuals with DLD showed a stronger association between the genetic disposition for psychiatric disorders and conduct problems than individuals without DLD. Given the high comorbidity between DLD and dyslexia(37), and the substantial genetic contribution to DLD(38), some overlap in social and behavioural outcomes may be expected. Although direct comparisons of results for DLD and dyslexia across these studies cannot be made, these findings provide broader support that genetic liability for neurodevelopmental language-relevant conditions may contribute to psychosocial outcomes later in life.

The strongest PGS association for dyslexia was observed for the amount of help received from colleagues on the work floor, where a higher genetic disposition for dyslexia predicted lower levels of perceived support (β=-0.48; *p*=0.007). Importantly, this measure specifically reflects experiences within the workplace rather than broader social interactions, highlighting the work environment as a potentially important context in which dyslexia-related social challenges arise. Previous studies examining dyslexia in the workplace relied primarily on phenotypic data and consistently reported difficulties and challenges. For example, dyslexia severity has been associated with lower work confidence (r=-0.30, *p*<0.01) and stronger negative emotions unique to dyslexia (r = 0.37, *p*<0.01)(30). In addition, a large meta-analysis that focussed on workplace experiences of individuals with dyslexia identified the largest effects for factors related to obtaining and maintaining employment, support from colleagues, and personal coping strategies(31). Cognitive challenges specific to the work floor for individuals with dyslexia have been investigated and confirmed as well; self-reported dyslexia traits were a significant positive predictor of workplace cognitive failures (β=0.247; *p*<0.001), even after controlling for ADHD symptoms, personality, well-being, and workplace characteristics^47^. A survey among employees with dyslexia suggested that employees with dyslexia particularly value workplace cultures characterized by awareness, inclusivity, and organizational support, highlighting the potential importance of supportive workplace environments for dyslexia-related challenges^48^. Our results might suggest that these wishes are currently not fully met. Given the central role of the workplace in adult life, these experiences may contribute to broader differences in well-being and life satisfaction, although this relationship was not evaluated in the present study.

Several limitations should be considered when interpreting these findings. First, the available GWAS summary statistics did not fully align with the available phenotypes in the Lifelines data. For example, no comparable GWAS was available for workplace social support or social self-efficacy, disabling the evaluation of the associations between dyslexia and these social traits with LDSC and MR, and also disabling investigation of relationships of PGS for these social traits with dyslexia outcomes. Second, although a range of sensitivity methods were applied, we cannot rule out violations of the MR assumptions or limited statistical power, particularly for the subjective well-being GWAS, for which only 8 SNPs met the 1E-6 significance threshold. However, decent instrument strength was supported by the F-statistics exceeding the conventional threshold (F > 10), and also the secondary MR analyses using a larger number of suggestive genetic instruments (*p*<1E-6) did not provide evidence for causality. Third, the PGSs used in this study only captured a proportion of the underlying genetic liability in the Lifelines cohort. For example, the PGS_dyslexia_ only explained 5.6% of the variance for dyslexia in the Lifelines cohort, while the captured h^2^_SNP_ in both the Lifelines (h^2^_snp_ = 13%) and the 23andMe sample (h^2^_SNP_ = 16%) was higher. Consequently, the PGS analyses with dyslexia and social traits captures only the component of genetic liability represented by the available genetic instruments and may therefore not fully reflect the genetic architecture underlying and relationships between these traits. Fourth, both dyslexia diagnoses and social-interactive measures were based on self-report, which might be less reliable compared to a clinical derived diagnoses or scores derived by an assessment. The prevalence for self-reported dyslexia diagnoses in the 23andMe sample (5%)(11) was slightly lower compared to the prevalence in the typical United States population (5.3-11.8%)(41). Prevalence of dyslexia in the Netherlands was estimated in 2005 at 3.6% for children at the end of primary school, and at 8.8% when children with severe reading- and spelling difficulties were included(42). These estimates align with the 8% dyslexia prevalence in the self-reported Lifelines sample. Furthermore, the 23andMe GWAS summary statistics on self-reported dyslexia showed high genetic correlations with quantitative measures of reading and spelling performance in the independent GWAS meta-analysis by GenLang(12), demonstrating that the signal captured by the 23andMe dyslexia self-report question is reliable. The social-based measures in the Lifelines data were self-reported as well, though all scores were derived from validated instruments, and were therefore considered as reliable and robust. Lastly, this study lacks ancestral diversity, as all analysis were conducted in individuals from European ancestry, which limits the generalizability of our findings to other populations(43). Follow-up studies should investigate whether these findings replicate in samples of non-European ancestry.

## Conclusion

This study provides empirical evidence that genetic disposition to dyslexia is associated with social interaction and well-being outcomes, while also offering insight into the potential directional relationship between dyslexia and overall well-being and life satisfaction. Together, these findings contribute to a broader understanding of the impact of dyslexia beyond literacy-related difficulties, highlighting its relevance for social and psychosocial functioning across the lifespan. We hope that this work encourages greater recognition of the challenges experienced by individuals with dyslexia and supports the development of targeted interventions and structural support systems that promote well-being and social participation throughout all stages of life.

## Methods

### Cohort data

For this study, phenotype and genotype data were obtained from Lifelines(17), a multi-disciplinary prospective population-based cohort study examining in a unique three-generation design the health and health-related behaviours of 167,729 persons living in the North of the Netherlands. It employs a broad range of investigative procedures in assessing the biomedical, socio-demographic, behavioural, physical and psychological factors which contribute to the health and disease of the general population, with a special focus on multi-morbidity and complex genetics. The data of the selected surveys on social well-being were collected between 2012-2019, and the “Speech, Language and Musicality” survey containing speech- and reading-based questions was administered in September and October of 2022. Questionnaires were distributed and completed in Dutch; English translations are used in the manuscript. The general Lifelines protocol has been approved by the UMCG Medical ethical committee under number 2007/152, and the “Speech, Language and Musicality” study was reviewed by the medical ethical committee of the Erasmus Medical Center (MEC-2022–0313). All participants provided informed consent for their participation in Lifelines (https://wiki.lifelines.nl/doku.php?id=informed_consent).

### Measures

#### Social interaction scales

The Lifelines study had several measures on social interactive experiences that were obtained via self-report surveys. For this study, we selected thirteen measures that are i) survey based, and ii) representative of various aspects of social well-being and/or social interaction (Table 2).

Results of the Social Production Function Instrument for Level of Well-being (SPF-IL)(44) questionnaire reflect the social aspects of well-being and individual experiences. Based on the theory of social production functions, the results that are captured reflect the state of physical well-being (comfort, stimulation) and social well-being (affection, confirmation, status). The latter three scales were used in this study. The Satisfaction with Life Scale (SWLS)(45) summarises an individual’s contentment with the overall course of their life and their current life status. The survey contains five distinct items, where scoring is captured on a seven-point agreeableness scale, that are combined into a single measure for life satisfaction. The experience of well-being specifically with regards to the workplace is captured within the Copenhagen Psychosocial Questionnaire (COPSOQ)(46). Within Lifelines, the second version was implemented, comprised of 21 items in total. The two scales representative of the social relationships experienced at work (social support from colleagues/social support from supervisor) were used in this study. The Self-Management Ability Scale (SMAS)(47) measures six self-management skills in social relationships and daily activities. The six skills are based on the theory of self-management of well-being, and reflect initiative-taking, self-efficacy beliefs, investment behaviour, positive mind-state, availability of resources, and variability of resources. Lastly, the Loneliness Scale (LS)(48) reflects the level of quantitative and qualitative social isolation an individual experiences. The survey captures social loneliness (lack of wider social network) and emotional loneliness (lack of intimate relationships) within a single measure.

#### Literacy-based measures

Reading- and writing ability was measured with three self-report items: whether the individual has dyslexia, whether the individual struggled with learning to read compared to their peers, and whether the individual’s writing has often been corrected by others. The survey-questions and answer-options per item are shown in Table 1.

### Phenotypic data cleaning and score coding

#### Exclusion criteria

Individuals who had suffered from a stroke (n=985) or who have been diagnosed with dementia (n=17) were excluded. Additionally, to ensure that non-native language would not influence an individual’s social interactive experience(49), individuals who were not born in the Netherlands were excluded as well (n=3,767).

#### Scoring of items and instruments

Traits representing reading-skills were coded as either binary or continuous (Table 1). Cases for individuals with dyslexia (question: “Do you have dyslexia?”) were defined as those who answered “Yes, I have been diagnosed” or “I presume I have it, but I have never been tested” (N_case_=2,639). Individuals who did not know (“I suppose not”) were excluded (N_exc_=3,175), and those who answered “No” were used as controls (N_control_=28,492). The remaining items (learning to read and spelling difficulty) were coded as continuous. To ensure consistency in scoring direction, scores were coded such that higher scores corresponded to more difficulty in reading and writing.

Coding of final scores for social-based surveys was done according to the established protocol for each respective questionnaire (SPF-IL(44); SWLS(45); COPSOQ(46); SMAS(47); LS(48)). For COPSOQ, we wanted to ensure that all individuals in the analyses were working with colleagues at the time of the survey. Therefore, we excluded participants from analyses that answered “no” to the question “Are you part of a working group at work?“ (N_exc_=15,396). There was no item in the questionnaire reporting on whether the individual had a supervisor/manager, thus we assumed that those who answered the supervisor-based items had a supervisor at the time of the survey. An overview of descriptives for the final scores can be found in Supplementary Table 1. Answer-options, scoring ranges, and sample sizes per trait are reported in Tables 1 and 2.

### Phenotypic correlation analyses

Correlation analyses were performed across all traits (reading and social) (Suppl. Table 2). Analyses were performed in R (version 4.2.2) using the Pearson-method. *P*-values were Bonferroni corrected to account for multiple testing.

### Psychological network analyses

In network analyses, psychological behaviour is conceptualized as a complex interplay of psychological and other components(50). The network consists of nodes representing observed variables, which are connected by edges representing statistical relationships. Estimation of the model is done using the Markov random field approach, where edges represent a full conditional association between two nodes after conditioning on all other nodes in the network. The resulting edges are defined as partial correlation coefficients. Mixed graphical models(21,22) are an extension of the Gaussian-Ising model and the class of exponential family Markov Random Fields, which allows for modelling of traits following binomial, continuous and categorical distributions within one network. Furthermore, it allows for correlations within and between different groups of variables and the model does not enforce connections across all nodes if a statistical relationship is absent.

Mixed graphical models were estimated using the mgm function(21) of the bootnet package(20) (version 1.6) in R (version 4.2.1) on the phenotypic correlation matrix. Following model estimation, bootstrap tests were performed in order to i) assess the edge-weight accuracy by construction of alternative 95% confidence-intervals (N_tests_ = 1,000), and ii) assess the node stability using a drop-case approach (N_tests_ = 1,000). Only models with a correlation stability coefficient (CS) of >0.7 were interpreted(20,51).

### Genotype data cleaning

Imputed genotyping data for this study were provided by UMCG Genetics Lifelines Initiative (UGLI) using data from two genotyping batches. The first batch (UGLI1, release 2) was genotyped using the Infinium Global Screening Array MultiEthnic Disease Version 1.0. Cleaned genotype data (filtering steps included INFO > 0.8, MAF>0.01, HWE *p≤*10^-6^, and call-rate>0.99) from 36,339 individuals and 548,029 markers were imputed with the Sanger imputation service using the Haplotype Reference Consortium reference panel (version r1.1), resulting in a total of 7,424,407 imputed SNPs. The second sample (UGLI2, release 2) was genotyped on the FinnGen Thermo Fisher Axiom custom array. The cleaned genotype data (the same filtering criteria used for UGLI1 were applied) of a total of 28,149 participants and 460,136 markers were imputed following the same protocol as used for UGLI1, resulting in a total of 7,474,700 imputed SNPs. For follow-up analyses, we used the principal components (PCs) provided by Lifelines, with PC scores generated separately for the genotyped individuals in UGLI1 and UGLI2. Further details about the genetic data provided by Lifelines can be found on their online wiki-page (https://wiki-lifelines.web.rug.nl/doku.php?id=ugli).

Follow-up preparation of the genetic files included additional filter steps (MAF>0.01, HWE *p≤*10^-4^, and imputation quality >0.8) and merging of the UGLI1 and 2 imputed genotype data, resulting in a total of 6,965,423 variants for 64,488 individuals. Filtering and merging were done with PLINK (v1.90b6.10).

### GCTA-GREML analyses

SNP-based heritability (h^2^_SNP_) was estimated using genome-wide complex trait analysis-genomic restricted maximum likelihood (GCTA-GREML) software(18) (version 1.93.2). The h^2^_SNP_ represents the proportion of phenotypic variance explained by the variation in genetic relatedness, captured by a genetic relationship matrix (GRM) estimated from genotyped SNPs. The genetic correlation represents the proportion of genetic variance that is shared between two traits, based on the GRM estimates.

Analyses were performed using the threshold-GRM approach(52) which allows related and unrelated individuals to be analysed jointly. A second GRM was constructed by setting relationship coefficient values (π) of <0.02 to 0, which enables simultaneous estimation of the pedigree heritability (h^2^_pedigree_) and heritability explained by genotyped SNPs (h^2^_SNP_). Modelling both GRMs together with GREML helps to distinguish SNP-associated genetic effects from additional familial resemblance, thereby reducing upward bias in SNP-heritability estimates due to shared environmental and pedigree-associated effects. Gender, genotype batch, age at time of assessment and the first 10 PCs were included as covariates.

### External GWAS summary statistics

Publicly available GWAS summary statistics representative of reading and social-interaction were selected for this study. We used the dyslexia GWAS summary statistics as the reading-based measure, which was based on self-report diagnoses of dyslexia (“*Have you been diagnosed with dyslexia?*”; “yes/no”) in the 23andMe cohort (n=1,138,870 with n_cases_=51,800)(11). We also selected five sets of social-interaction and well-being based GWAS summary statistics. The subjective well-being GWAS summary statistics came from a meta-analysis study of items evaluating levels of happiness(16). The original meta-analysis consisted of 59 participating cohorts with a total sample size of n=298,420. However, the publicly available data from that well-being study do not contain the 23andMe cohort. Furthermore, as the GWAS meta-analysis also included the Lifelines cohort, we received GWAS results excluding Lifelines.

The final total sample size included 192,994 individuals. All three loneliness-based measures (sociability, loneliness, and social isolation) were constructed from composite-scores in UK Biobank: the sociability GWAS summary statistic was based on (1) frequency of family/friends visits, (2) number and type of visited social venues, (3) worrying about social embarrassment, and (4) loneliness experience, with 342,461 participating individuals(14). The loneliness GWAS summary statistics were constructed from (1) frequency of feeling lonely, (2) number of individuals in the household, (3) frequency of family/friends visits, and (4) the ability to confide in someone, with a total of 452,302 individuals(13). GWAS summary statistics for social isolation were derived from (1) frequency of family/friends visits, (2) ability to confide in someone, and (3) number of social activities per week, with a sample size of 449,609(15) individuals. Lastly, the social trust GWAS summary statistics were based on a composite score of (1) trustworthiness of other people, (2) if others take advantage of you, and (3) if others display altruistic behaviour(25). This latter study was conducted in a Danish cohort with a sample size of 33,882 individuals. A complete overview of the GWAS summary statistics can be found in Suppl. Table 5.

### LDSC analyses

SNP heritability of and genetic correlations across GWAS summary statistics were estimated with Linkage Disequilibrium score regression (LDSC)(23,24). LDSC software (v1.0.1) was used to munge summary statistics, followed by estimation of the heritability and genetic correlations. LD scores were based on the phase 3 1000 Genomes Project European reference genome panel, and analyses were restricted to a pre-defined set of HapMap 3 SNPs.

### Power prediction for cross-trait PGS analyses

Power calculations were performed in order to estimate the expected predictive accuracy of PGS derived from selected GWAS summary statistics in Lifelines. First, the maximum possible prediction accuracy *R^2^* for the summary statistic was estimated by fitting the variant effect sizes using a best linear unbiased prediction(26),

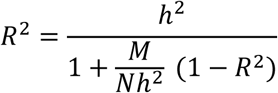

where *h^2^* corresponds to the summary statistic-based h^2^_SNP_ as estimated with LDSC, *N* represents the GWAS sample size, and *M* represents the number of independent common variants that inform the PGS (for these analyses, *M* was based on the number of independent common SNPs that are present on a standard genotyping array for European ancestry(53), which is *M* = 60,000).

The maximum possible *R^2^* value for a within-trait predictor-outcome trait combination (e.g. dyslexia-dyslexia) was used to further inform *R^2^* for cross-trait predictor-outcome trait combinations (e.g. dyslexia-life satisfaction). We multiplied the maximum *R^2^* of the predictor GWAS with the genetic correlation (r_g_) between dyslexia and social traits, as estimated with LDSC, to account for the level of shared genetic variance in the cross-trait PGS analysis.

Then, we used a range of *R^2^* values to estimate the power for the cross-trait PGS analyses using simulations. First, a multivariate normal distribution was generated with mean 0 and a correlation parameter r defined as the square root of the adjusted R^2^, with the sample size equal to that of the outcome trait. Subsequently, a regression was conducted across the constructed multivariate data. Simulations were repeated 10,000 times, where the power was estimated based on the frequency (percentage) of successful predictions (where *p<*0.0125).

### PGS analyses

PGS analyses were conducted to further investigate the genetic overlap between reading- and social-based traits. These analyses were conducted among unrelated individuals; related individuals were excluded based on a GRM threshold of π>0.05 using GCTA (version 1.93.2), resulting in 23,160 individuals that were included in the analyses. The cleaned and imputed UGLI1 and UGLI2 genotyping data were used, with additional filtering steps (INFO > 0.8; MAF>0.01; HWE *p≤*10^-6^; variant genotype-missingness >0.01; sample genotype-missingness >0.01; ambiguous SNPs) (PLINK v1.90b6.10), leaving a total of 4,308,372 variants for PGS analyses. (No individuals were removed with the additional filtering steps).

PGSs were estimated following the PRS continuous shrinkage (PRS-CS) framework(54), where SNP weights are adjusted for LD structures in the sample. The resulting SNP effect sizes account for LD patterns in the target sample, which improves predictive accuracy of the PGS. All β-effect sizes of the selected summary statistics were adjusted with the 1000 genomes reference panel based only on the available SNPs in the Lifelines genotype data. β-effect size adjustment was done with PRS-CS software (v1.1.0-10-gf86d5c7), after which the participant’s PGS were estimated with PLINK (v1.90b6.10).

Regression analyses were conducted in R (v4.2.2) using the glm() function for binary outcomes, and the lm() function for continuous outcomes. First, the null model was constructed with the age at the time of assessment, the first five PC’s, and gender as predictors, and the z-standardized phenotype score for dyslexia or social-based trait as outcome. The PGS-model included the z-standardized PGS as predictor as well. For the regressions with a binary outcome, McFadden R^2^ values were calculated based on the fitted model’s log likelihood values, and odds ratios were derived from the regression results. For the regressions with a continuous outcome, incremental R^2^ values were estimated by deducting the null-model r^2^ from the PGS-model r^2^, and β-estimates were derived from the PGS-model.

### Mendelian randomization analyses

The causal effect of exposure-associated SNPs on the outcome was estimated using the Mendelian Randomization (MR) framework, which uses genetic variants as instrumental variables to study the causal effect of an exposure (the instrumental SNPs associated with the exposure trait) on an outcome. Within the MR framework, three core assumptions must be met for the results to provide reliable evidence of causality(55,56). First, the instrumental variants must be robustly associated with the exposure. Therefore, only SNPs reaching genome-wide significance (*p* <5E-8) in the exposure GWAS were selected as instruments. Second, the instrumental variants must be independent of potential confounders of the exposure–outcome relationship. Third, the instrumental variants must not directly affect the outcome, except through their effect on the exposure. Further details on MR methods and additional/other assumptions are outlined in Lukas et al. (2025)(57) and Burgess et al. (2023)(55).

Prior to MR analyses, SNP identifiers (rsIDs) of summary statistics were updated based on chromosome and base-pair position, and aligned against HRC.r1 (build GRCh37). Lead SNPs were subsequently selected (PLINK v1.9b6) based on their association *p*-value (<5E-8 for the genome-wide significant SNPs, and <1E-6 for genome-wide suggestive SNPs) and clumped with an r^2^ of 0.01 and a window-size of 10 kb. The resulting lead SNPs were used as instrumental variables in the MR analyses. After selection of SNPs present in both the exposure- and outcome-GWAS summary statistics, data were harmonized based on alleles and their corresponding frequencies.

MR analyses were conducted using the TwoSampleMR(28,29) (v0.6.11) package in R (v4.2.1). Instrument strength towards the exposure was assessed with the F-statistic(58), retaining only those with F>10 to reduce bias from weak instruments. Next, causal effects were inferred from the SNP–exposure and SNP– outcome associations combined. The inverse-variance weighted (IVW) method was used as the primary method, as recommended by the MR guidelines(55), which combines SNP-specific Wald ratios and provides an unbiased estimate when all instrumental variable assumptions are met or when pleiotropy is balanced. Additional sensitivity analyses were performed to assess robustness of the results and potential MR violations. These included the MR Egger regression, which allows for directional pleiotropy by estimating an intercept term, the Weighted median estimator, which provides a consistent estimate if at least 50% of the weight comes from valid instruments, and the Weighted mode and Simple mode, which identify the most common causal effect estimate across SNPs under different clustering assumptions.

Follow-up sensitivity analyses for heterogeneity and pleiotropy were further evaluated using regression diagnostics and graphical inspection of SNP-specific effects, including funnel and scatter plots. Influence analyses were performed using leave-one-out analyses to assess whether results were driven by individual SNPs, and SNP-specific Wald ratio estimates were examined in relation to their standard errors to identify potential outliers.

## Author contributions

DA and EE conceptualized and designed the study. DA performed the analyses, interpreted the data, created figures, and wrote the draft of the manuscript. EE and SEF supervised the research and critically reviewed the manuscript. RV provided guidance and feedback on the mendelian randomization analysis.

## Data Availability

Researchers can apply to use the Lifelines data used in this study. More information about how to request Lifelines data and the conditions of use can be found on their website.

## Acknowledgements

DA, EE, and SEF were financially supported by the Max Planck Society. The Lifelines initiative has been made possible by subsidy from the Dutch Ministry of Health, Welfare and Sport, the Dutch Ministry of Economic Affairs, the University Medical Center Groningen (UMCG), University of Groningen and the Provinces in the North of the Netherlands (Drenthe, Friesland, Groningen).

