## Supplemental Figures for "Dyslexia extends beyond reading difficulties: genetic links with social functioning and well-being"

### Supplementary material

Supplementary Figure 1: $h^2_{SNP}$ estimates across reading- and social-based traits categorized by group, estimated in Lifelines data. Error bars show 95% confidence intervals.

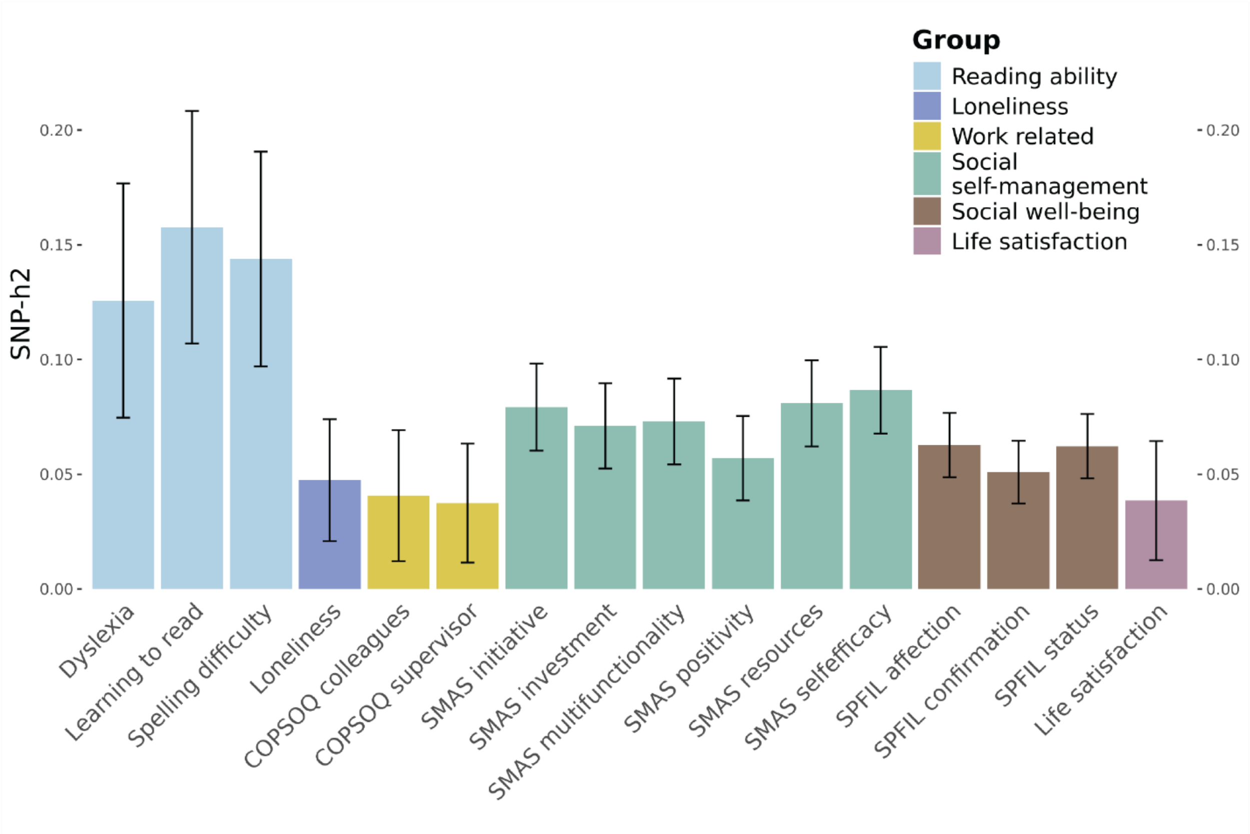

Supplementary Figure 2: Power predictions for PGS analyses with dyslexia as predictor. Maximum possible $R^2$ (variance explained by PGS) is shown, based on prediction of similar trait (e.g. dyslexia predicting dyslexia). The plots display the amount of estimated power (%) for various $R^2$ values. Vertical lines display the associated $R^2$ value for various $r_g$ values between predictor and outcome (e.g. dyslexia predicting loneliness).

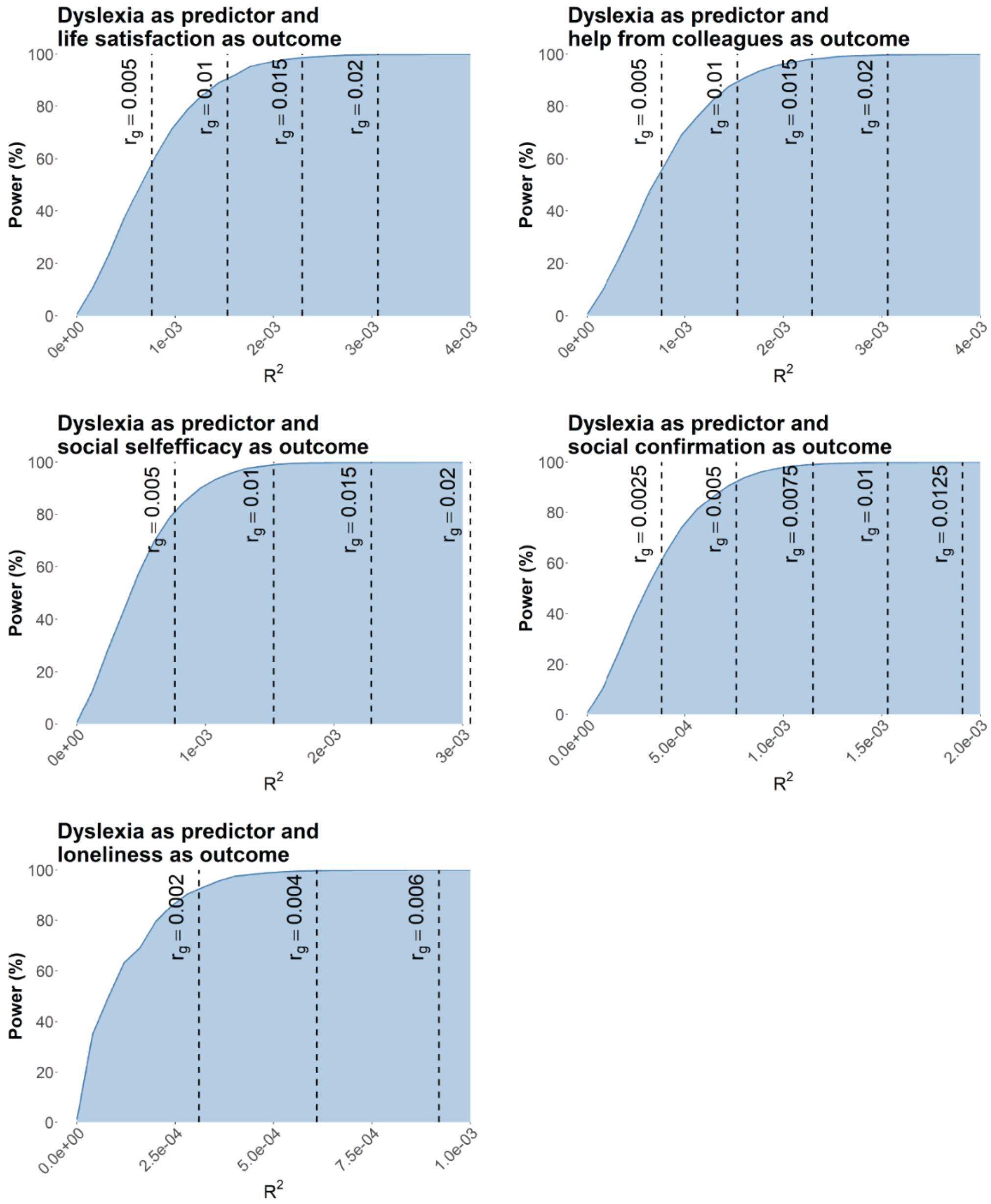

Supplementary Figure 3: (**A**) Power predictions for PGS analyses with social-based trait as predictor. Maximum possible $R^2$ (variance explained by PGS) is shown, based on prediction of similar trait (e.g. dyslexia predicting dyslexia). The plots display the amount of estimated power (%) for various $R^2$ values. Vertical lines display the associated $R^2$ value for various $r_g$ values between predictor and outcome (e.g. dyslexia predicting loneliness). (**B**) LDSC-based genetic correlations between dyslexia and the five social-based traits. *= significant after Bonferroni correction, with $p_{bonferroni}<0.05/5=0.01$.

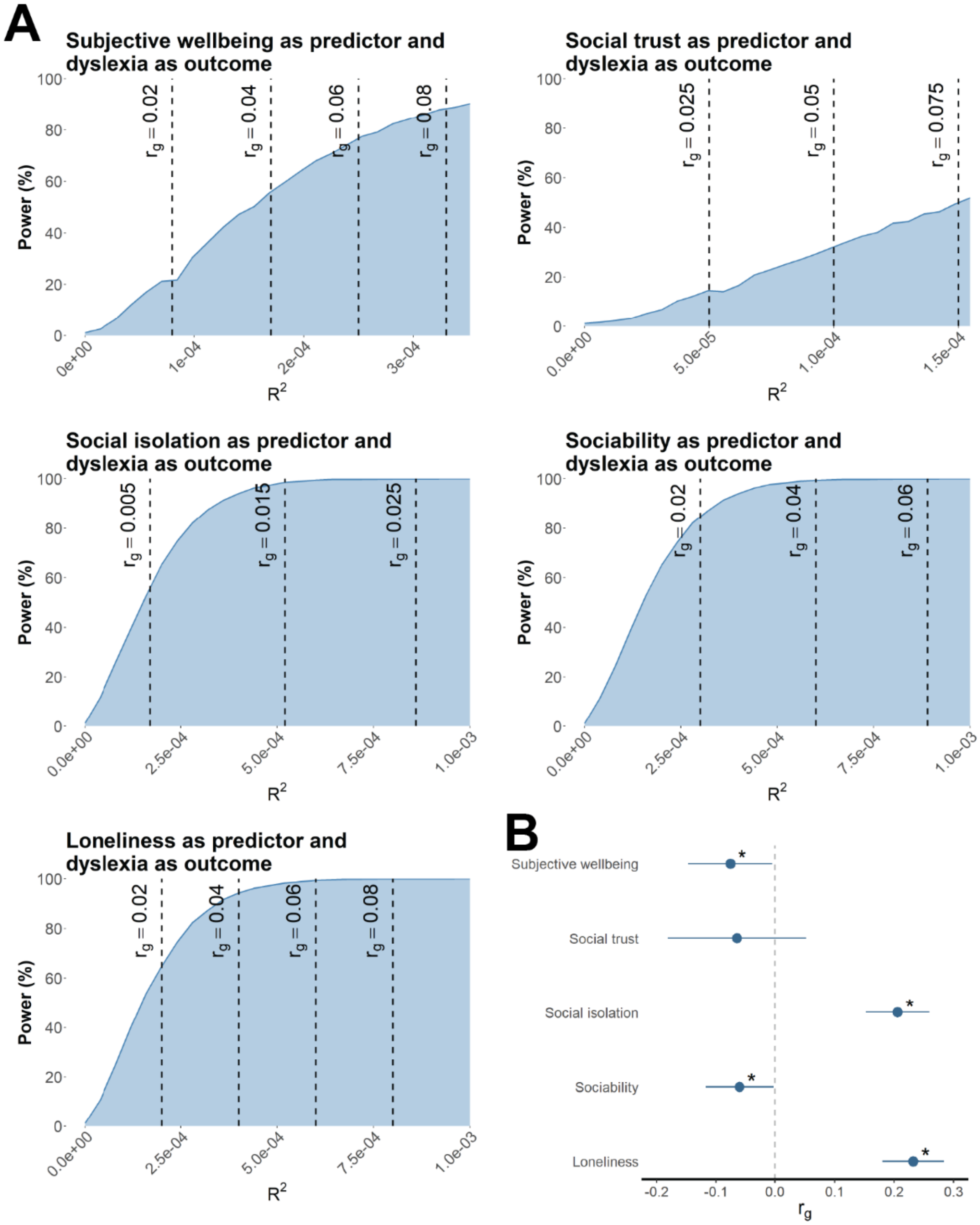

Supplementary Figure 4: PGS prediction of (**A**) dyslexia and (**B**) social-based measures. The explained variance is shown for either continuous (Incremental R2) or binary (McFadden R2) outcomes (*= significant after Bonferroni correction, with $p_{Bonferroni}$=0.05/20 tests=2.5E-3), followed by effect sizes, for either continuous (beta [β]) or binary (odds-ratio [OR]) outcomes, with error bars representative of 95% confidence intervals. Predictor GWAS summary statistics: Dyslexia by Doust et al. (2022)[1]; Loneliness by Day et al. (2018)[2]; Sociability by Bralten et al. (2021)[3]; Social isolation by Socrates et al. (2024)[4]; Social trust by Sequeros et al. (2024)[5]; Subjective well-being by Okbay et al. (2016)[6].

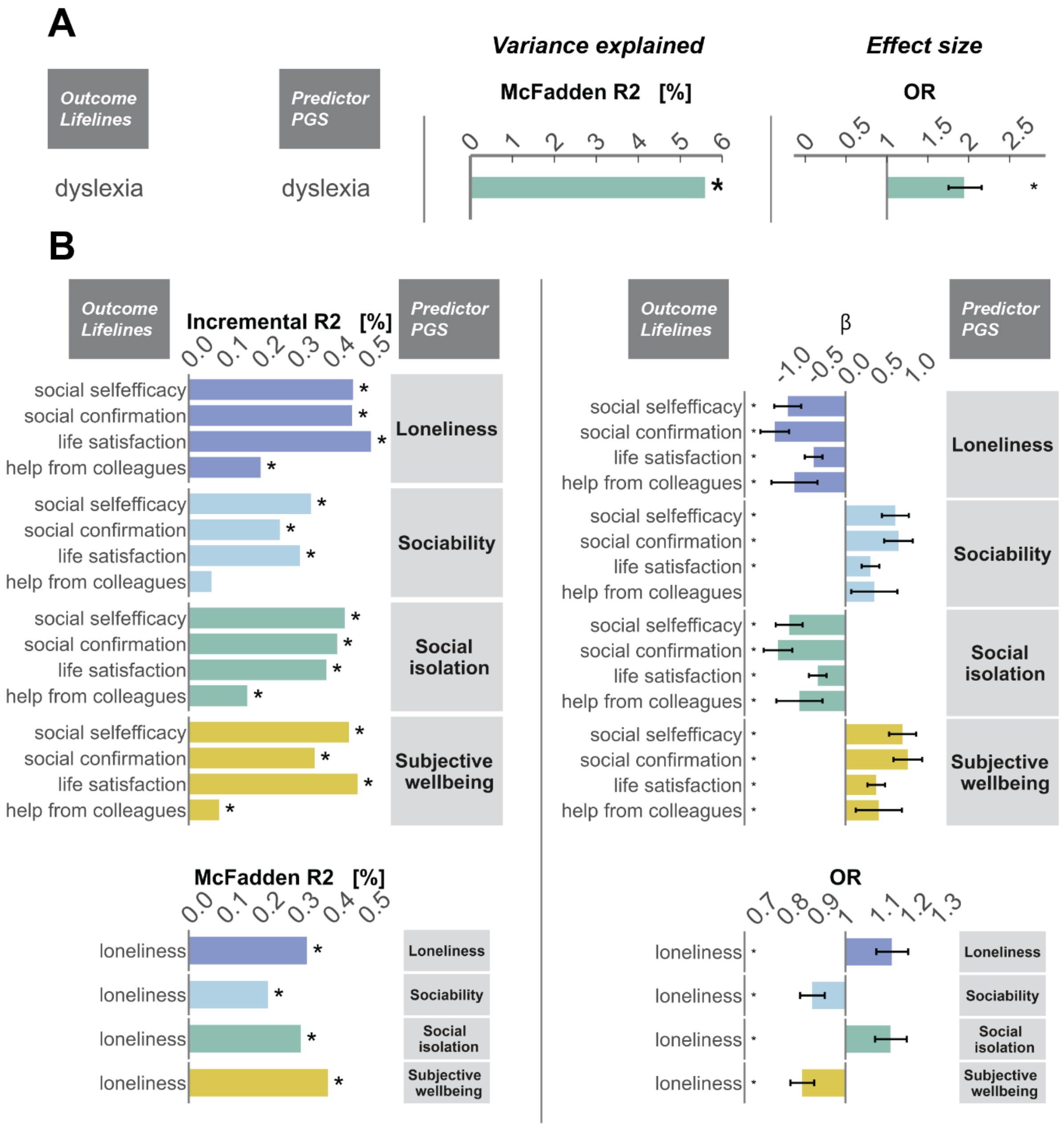

Supplementary Figure 5: (**A**) Distribution of cases/controls across dyslexia within the Lifelines cohort per age-group. Percentage of dyslexia-cases is shown on the y-axis. Only age groups with >10 individuals are shown (remaining age groups [$n_{cases}$/$n_{controls}$]; 88 [0/3]; 89 [0/5]; 90 [0/2]). (**B**) Prediction of reading-based outcomes with PGS for dyslexia and social isolation in Lifelines cohort. Explained variance for binary (McFadden $R^2$) and continuous (Incremental $R^2$) outcomes is reported, as well as the effect size, again for both binary (OR) and continuous (β) outcomes, with estimated 95% confidence intervals. *=significant after Bonferroni correction for five tests, with $p_{Bonferroni}$=0.05/5 tests=0.01. **= Bars not shown, as incremental $R^2$ was small (learning to read = 4.9E-3%; spelling difficulty= 6.0E-3%).

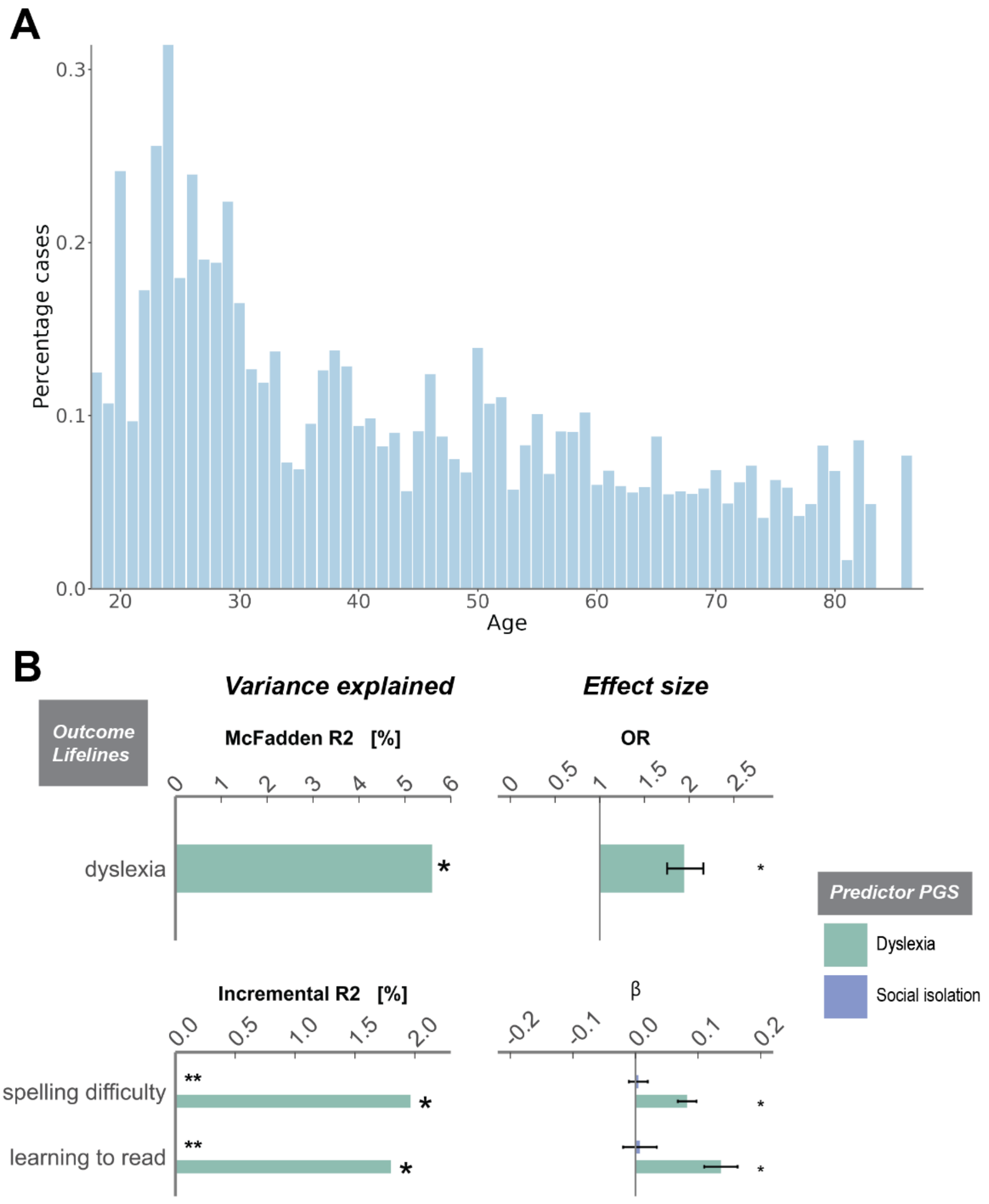

Supplementary Figure 6: Mendelian Randomization (MR) analyses based on GWAS summary statistics for dyslexia[1] and social traits[2–4,6]. Panels show (**A**) the primary analysis with lead SNPs selected with a GWAS *p*<5E-8 (circles), and (**B**) a secondary analysis with suggestive SNPs with GWAS *p*<1E-6 (squares). Error bars represent 95% confidence intervals. Left panel shows the MR estimates with dyslexia as exposure variable, and social traits as outcome variable. The right-panel shows the MR estimates with the social traits as exposure variables and dyslexia as the outcome, with black dotted lines highlighting the range of-0.2>β>0.2. Black filled circles and squares represent results with *p*<0.05, and ***** represent *p* after Bonferroni correction (*p* < 0.05/4 tests= 1.3E-2). Right-panel notes: *95%CI are cut-off at-2 and 2. **Exposure GWASs that did not provide SNPs with *p*<5E-8 are highlighted with X. IVW = Inverse variance weighted; mod = mode; med = median.

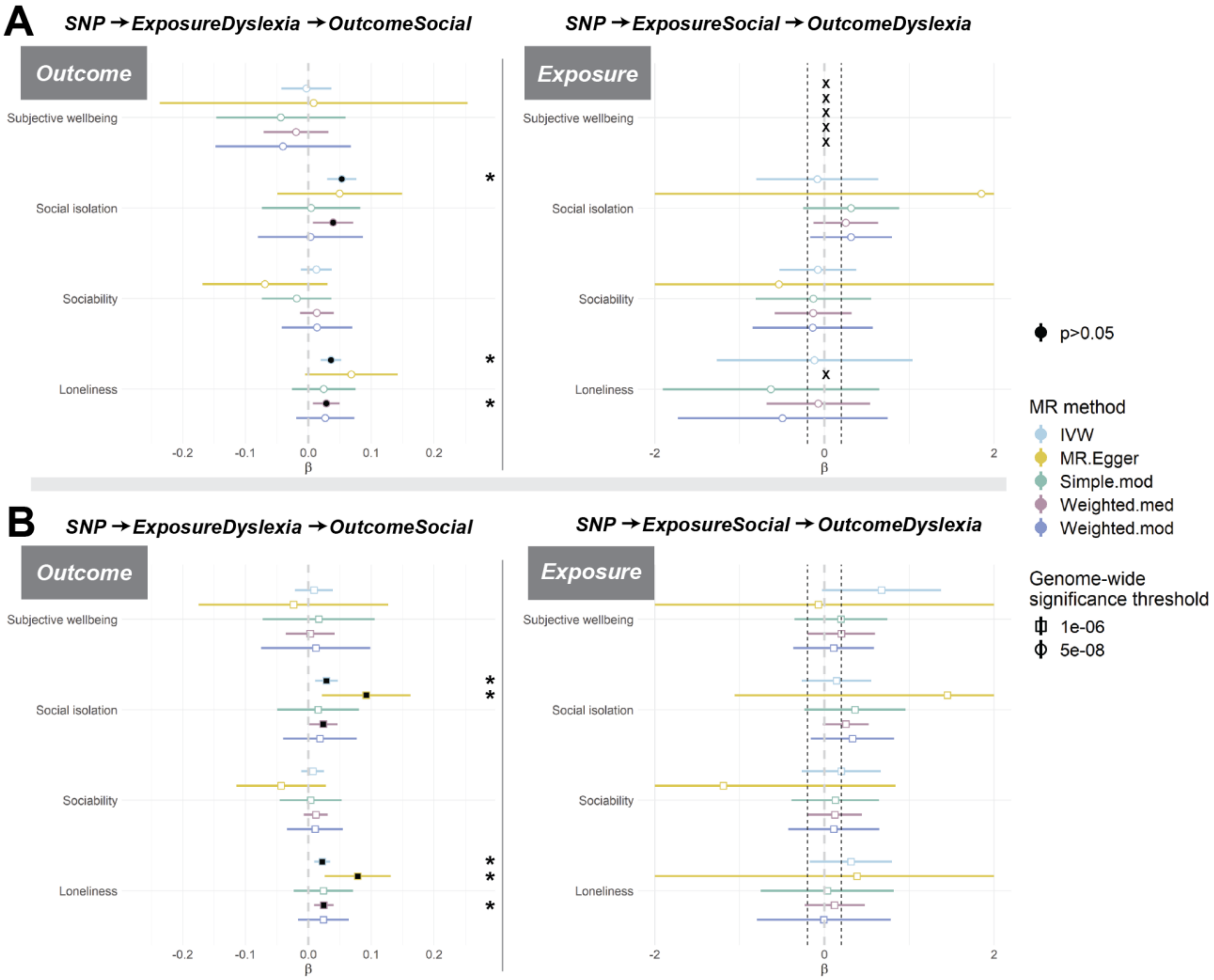

Supplementary Figure 7: Leave-one-out sensitivity analyses for the causal effect of dyslexia on social traits. The x-axis reports the IVW estimate for every SNP when excluded from analysis (y-axis). The red line at the bottom of the plot defines the estimated IVW value for the analysis with all instrumental SNPs. Bars represent 95% confidence intervals. IVW = Inverse variance weighted

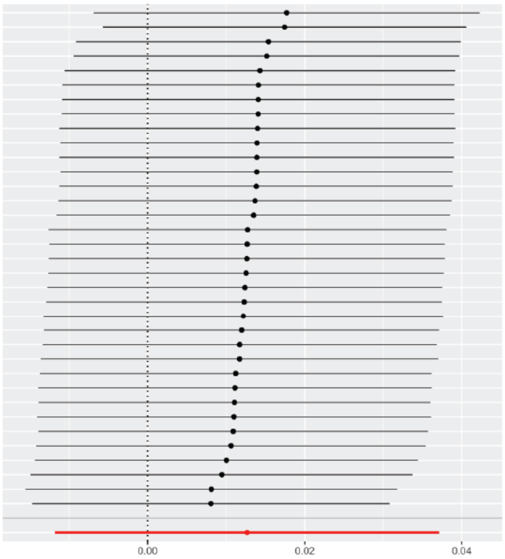


MR leave-one-out sensitivity analyses for dyslexia on sociability

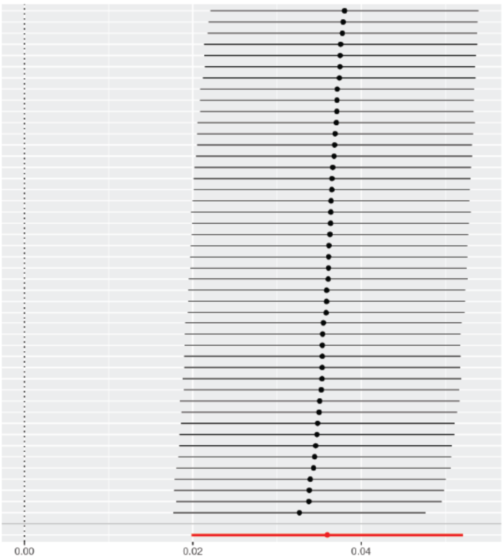


MR leave-one-out sensitivity analyses for dyslexia on loneliness

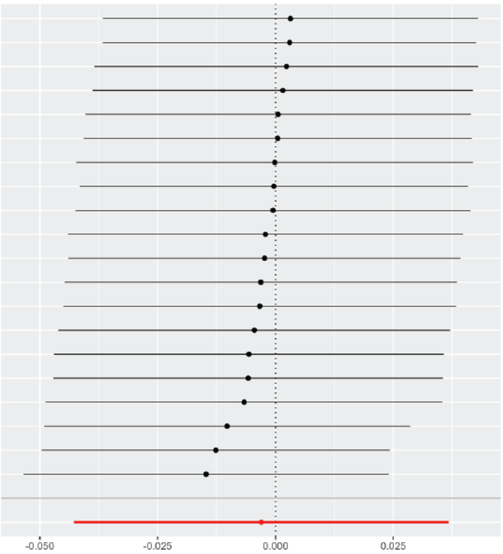


MR leave-one-out sensitivity analyses for dyslexia on subjective well-being

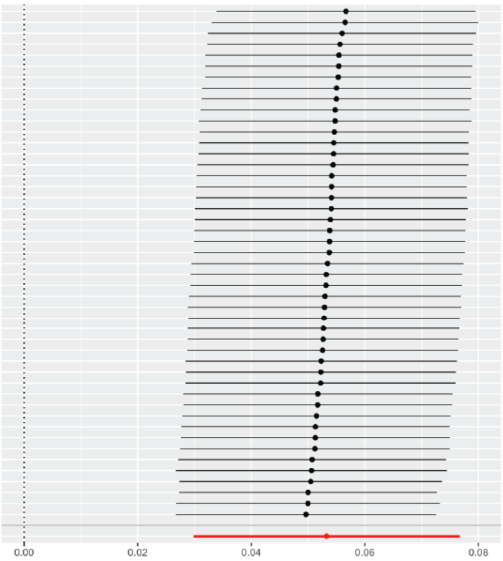


MR leave-one-out sensitivity analyses for dyslexia on social isolation
